# New IPECAD open-source model framework for the health technology assessment of early Alzheimer’s disease treatment: development and use cases

**DOI:** 10.1101/2024.04.05.24305373

**Authors:** Ron Handels, William L. Herring, Sabine Grimm, Anders Sköldunger, Bengt Winblad, Anders Wimo, Linus Jönsson

## Abstract

**Objectives:** Market access and reimbursement decisions for new Alzheimer’s disease (AD) treatments are informed by economic evaluations. An open-source model with intuitive structure for model cross-validation can support the transparency and credibility of such evaluations. We describe the new IPECAD open-source model framework (version 2) for the health-economic evaluation of early AD treatment and use it for cross-validation and addressing uncertainty.

**Methods:** A cohort state transition model using a categorized composite domain (cognition and function) was developed by replicating an existing reference model and testing it for internal validity. Then, features of existing “ICER” and “AD-ACE” models assessing lecanemab treatment were implemented for model cross-validation. Additional uncertainty scenarios were performed on choice of efficacy outcome from trial, natural disease progression, treatment effect waning and stopping rules, and other methodological choices. The model is available open-source as R code, spreadsheet and web-based version via https://github.com/ronhandels/IPECAD.

**Results:** In the IPECAD model incremental life years, QALY gains and cost savings were 21-31% smaller compared to the ICER model and 36-56% smaller compared to the AD-ACE model. IPECAD model results were particularly sensitive to assumptions on treatment effect waning and stopping rules and choice of efficacy outcome from trial.

**Conclusions:** We demonstrated the ability of a new IPECAD opens-source model framework for researchers and decision-makers to cross-validate other (HTA submission) models and perform additional uncertainty analyses, setting an example for open science in AD decision modeling and supporting important reimbursement decisions.

## INTRODUCTION

Market access and reimbursement decisions for new Alzheimer’s disease (AD) treatments are informed by economic evaluations. Recently, AD drugs lecanemab (1) and donanemab (2) have been tested in phase 3 randomized trials. They targeted persons with mild cognitive impairment (MCI) or mild dementia who had abnormal levels of amyloid pathology, a biological hallmark of AD hypothesized to cause dementia. The trial results showed significant reductions of amyloid pathology and slower decline on clinical scales measuring cognition and function. Other drugs are under development and being tested in phase 3 trials.

The European Medicines Agency is currently assessing the quality, safety and efficacy of lecanemab with a planned decision for market authorization early 2024 (3). The National Institute for Health and Care Excellence (NICE) in the United Kingdom (UK) is currently appraising the clinical and cost-effectiveness of lecanemab with a planned publication in July 2024 (4). NICE, based on a stakeholder workshop (5), identified key issues in the assessment of new drugs for treating AD. These included lacking evidence on the validity of surrogate endpoints, meaningfulness of the clinical outcomes and limited understanding of natural disease progression. Other issues are the potentially large budget impact, the role of new diagnostic pathways and risk of inequalities by disparities in access to treatment (6). NICE highlighted the importance of transparency and credibility of decision-analytic economic models, a known issue in policy making (7). The use of open-source models for healthcare decision making has been considered very important by most responders of a survey among stakeholders in academia, industry and HTA agency (8).

In their review of transparency in decision modeling, Sampson et al. (9) found that transparency is manifested through open-source modeling in addition to collaboration, peer review, reference models, reporting standards and model registration. In addition, the concept of model cross-validation has been defined as “examining different models that address the same problem and comparing their results” in good research practice guidelines (10) and has been argued it could increase confidence in models if similar results are observed.

In 2019 the IPECAD open-source model (version 1) has been developed (11). It has been re-used outside its developing team (12, 13), used for cross-comparing a single model (14, 15) and used for cross-comparison among multiple models (16, 17). Following recommendations, it reflected disease progression by multiple domains of cognition, function and behavior (18–20). However, we experienced difficulties implementing a treatment effect comparable to those observed for new AD treatments, requiring assumptions on dependencies between domains and calibration, which we think limited model transparency.

For the appraisal of current and possible future AD drugs, we argue that an open-source model with intuitive model structure that is easy to use for model cross-validation is urgently needed to support transparency and credibility of new AD drug cost-effectiveness assessments. Therefore, we describe the new IPECAD open-source model framework (version 2) for the health-economic evaluation of early AD treatment and aim to apply it in 3 use cases for AD lecanemab treatment: 1) cross-validating an existing model with a similar structure (ICER (21)), 2) cross-validating an existing model with a more complex structure (AD-ACE (22)) and 3) assessing additional uncertainty scenarios.

We intend the use cases to act as an example of how the new IPECAD open-source model framework could support the cross-validation of a model submitted for appraisal to a reimbursement agency and support addressing uncertainty. We note a detailed evaluation of lecanemab is outside the scope of this study.

## METHODS

The new IPECAD open-source model framework was developed based on an existing cohort state-transition AD disease progression reference model assessing the potential health-economic impact of a hypothetical treatment in MCI due to AD (23). This model consists of states MCI and mild, moderate and severe dementia and death. We used original non-rounded transition probability input estimates available from the authors (AW and RH). The replicated model produced the same model outcomes in terms of mean person-years per state and alive when rounded to 2 decimal points, except for person-years in severe dementia after 40 years which had an absolute deviation of 0.01 person-years. We judged these outcomes as a sufficient reflection of internal validity. Next, we implemented new features for care setting, treatment stopping rules and treatment effect waning (i.e., decreasing of a treatment’s effect).

### MODEL RATIONALE, DESCRIPTION, ANALYTICS AND ASSUMPTIONS

We opted for a simple, commonly used model structure with transitions between states and a 1-year cycle length to serve transparency and credibility to its users. **Figure 1** shows the model structure detailing disease severity states as MCI and mild, moderate and severe dementia. It includes separate states for care setting and treatment status in MCI and mild dementia (i.e., on vs. off treatment) to allow various assumptions on treatment discontinuation and treatment effect waning.

**Figure 1:**
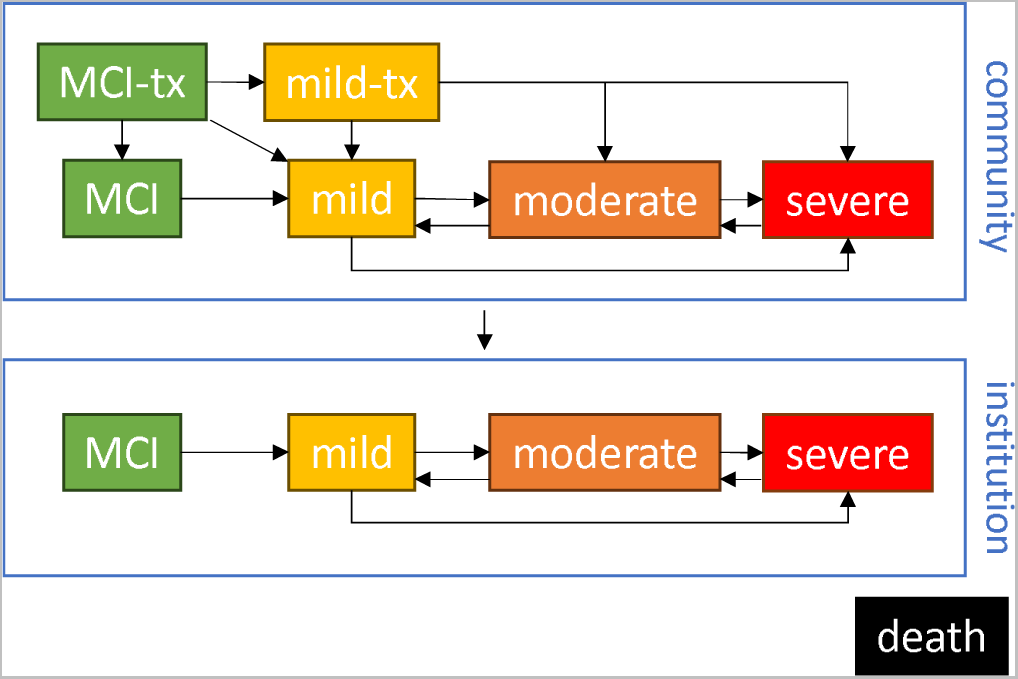
Graphical representation of new IPECAD open-source model framework. A limited number of all transitions (detailed in text) are shown. Abbreviations: MCI, mild cognitive impairment; tx, on treatment.

The characteristics of the starting population include age, sex distribution and proportion in MCI and mild dementia (latter in off-treatment state for standard of care strategy and on-treatment state for intervention strategy).

Transitions among MCI and mild, moderate and severe dementia states reflect disease progression. Forward transitions (i.e., to more severe disease states) are allowed between all disease severity states. Backward transitions are allowed to all disease severity states except for transitions from moderate or severe dementia to MCI as these were not observed (24). Transitions are assumed to be time-independent (i.e., Markov assumption). This aligns with age not being a significant predictor (23), being a relatively small factor (25) or being adjusted for (24), but differs with progression dependent on time in state (26). Probabilities for remaining in the same state are calculated as 1 minus the probabilities for transitioning to other states.

Transitions from community to institution reflect changes in care setting. All disease severity states are duplicated for community and institution settings. The probability of transitioning from the community setting to the institution setting is conditional on disease severity, unidirectional (i.e., only from community to institution with no back-transition) and time independent.

Transitions to death can occur from any disease severity state. Mortality related to AD natural progression is assumed to be multiplicative to general population mortality. This is operationalized by multiplying the probabilities of death from a general population age- and sex-specific life table with the relative risk of death in each disease severity state. Due to age-specific mortality, the starting population reflects a single specific age and not an age range. Transitions between states other than death are assumed conditional on remaining alive during the cycle in question. We note transitions to death indirectly reflect time dependency in the form of age-specific mortality.

Transitions from on- to off-treatment reflect treatment discontinuation. Disease states for MCI and mild dementia in the community setting only are duplicated for on- and off-treatment. Disease states for MCI and mild dementia in the institution setting and for moderate and severe dementia in both settings reflect off-treatment. The probability from on- to off-treatment is independent of health state and unidirectional. This setup reflects the assumptions that treatment is discontinued at moderate dementia and that treatment is never provided in an institution care setting. Treatment discontinuation is operationalized as time-dependent probabilities of transitioning from on- to off-treatment. These are set over an initial period (e.g., due to side effects), a later period (e.g., due to health events interfering with receiving treatment) and a maximum duration.

Treatment effect relative to a control strategy is implemented as a relative risk multiplied to each of the transition probabilities from MCI to mild, moderate and severe dementia and from mild dementia to moderate and severe dementia. Specific relative risks can be set per transition between disease states and separately for on and off treatment. This latter feature facilitates consideration of a remaining treatment effect even after treatment is no longer provided.

Treatment effect waning is operationalized by a waning factor by which the treatment relative risk is raised (e.g., a treatment relative risk of 0.70 and 0.15 waning gives 0.70^((1-0.15)^cycle)). This can be specified separately for transitions on-treatment and, if applicable, transitions off-treatment.

Utilities (separate for patient and informal caregiver) and costs by health, social and informal care sector are specified per disease severity and care setting state (community or institution). Treatment costs are set to all on-treatment states. Caregiver utility was set to zero after patient death. No default setting is chosen and for this study aligns with the United States (US) setting for the cross-validation use cases.

A state trace (i.e., proportion of patients in each state at each timepoint) is calculated. A half-cycle correction is operationalized by taking the mean of each 2 adjacent timepoints for life years, QALYs and cost outcomes in each cycle. Cycle 1 is the period between time 0 and time 1, with starting age applied to cycle 1. The model allows for discounting life years, QALYs and costs. Inputs are provided per cycle to facilitate a different cycle length (detailed below). Multiplication between relative risks and transition probabilities are done on rate scale (27).

See **supplemental material Table S1** column 1 and 2 for a list of all input parameters of the new IPECAD open-source model framework and a short description.

### USE CASE 1: CROSS-VALIDATE EXISTING ICER MODEL

The first use case is the cross-validation of a model developed by the Institute for Clinical and Economic Review (ICER) in the US. ICER is an independent non-profit research organization that evaluates medical evidence and convenes public deliberative bodies to help stakeholders interpret and apply evidence to improve patient outcomes and control costs (icer.org). They developed a cohort state-transition model with AD disease severity states to estimate the lifetime cost effectiveness of lecanemab in comparison with the standard of care (21, 28). Its development is linked to an earlier assessment of aducanumab arguing the model conceptualization (29, 30) and its basic model structure dates back to another AD state transition model developed earlier (31). We sought to replicate all features of the ICER model given the similarities with the IPECAD model structure.

### USE CASE 2: CROSS-VALIDATE EXISTING AD-ACE MODEL

The second use case is the cross-validation of a model published by Tahami Monfared et al. (22) which was funded by Eisai, the sponsor of the lecanemab phase 3 trial. They used the Alzheimer’s Disease ACE Simulator (AD-ACE) model (32) which uses a patient-level microsimulation approach capturing domains for AD pathology (e.g., beta-amyloid biomarkers), cognition, function, behavior and dependency. Its development is linked to earlier studies from Tahami Monfared et al. (33, 34), Kansal et al. (32) and Getsios et al. (35). We sought to mirror their input parameters for the cross-validation, given the different model type does not allow for replication.

### USE CASE 3: ADDITIONAL UNCERTAINTY SCENARIOS

To show the potential of the new IPECAD open-source model framework we performed univariate sensitivity analyses on choice of efficacy outcome from trial, natural disease progression, treatment effect extrapolation (i.e., waning) and stopping rules, and other methodological choices. These factors were chosen as they have been shown to have (or are anticipated to have) a large impact on health-economic outcomes (5, 16, 17), are sometimes complex to implement (as compared to parameter uncertainty) or to our knowledge have not been extensively tested in earlier studies. These sensitivity analyses were performed using the lecanemab scenario as implemented by ICER. See **Table 2** for details on these scenarios.

### USING THE MODEL

The new IPECAD open-source model framework is available in multiple formats (R code, spreadsheet and web-based). The R version uses base R combined with the dampack package to facilitate model outcome visualization and sensitivity analysis. We followed a coding guideline (36) commenting the R code, using object prefixes and following a similar structure as R dampack vignettes (37, 38). The spreadsheet version (in open document spreadsheet format) and web-based (R shiny) version does not contain probabilistic sensitivity analysis functionality. The model is hosted on GitHub (https://github.com/ronhandels/IPECAD) for its features of version control and collaboration.

Most model features are optional and can for example be turned off by setting them to 0 (e.g., transition probabilities, utilities or costs) or 1 (e.g., treatment relative risks). Experienced users may add or adjust model features to suit their research objectives.

## RESULTS

### USE CASE 1: CROSS-VALIDATE EXISTING ICER MODEL

Almost all features of the ICER model were implemented. However, starting in institution care setting and age-specific utilities and costs could not be implemented and some features were open for interpretation. See **supplemental material Table S1** and its table notes for a list of all input parameters and short description.

Compared to the ICER the IPECAD model mean per person outcomes showed higher life expectancy (+0.38 year), similar QALYs and lower costs (-$185,000) in the standard of care strategy. Incremental (intervention minus standard of care strategy) life years, QALY gains and cost savings were smaller (21-31% smaller) (see **Table 1**). The incremental cost-effectiveness ratio was somewhat larger ($272,000 instead of $236,000 per QALY gain).

**Table 1:** Cross-validation ICER and AD-ACE models; reported base case discounted model outcomes from ICER, AD-ACE and the IPECAD model outcomes from their cross-validation.

|  | ICER model |  |  | AD-ACE model* |  |  |
| --- | --- | --- | --- | --- | --- | --- |
|  | Original model | IPECAD cross-validation | Difference | Original model | IPECAD cross-validation | Difference |
| *Life years* |  |  |  |  |  |  |
| Standard of care | 5.77 | 6.15 | 0.38 | 5.61 | 6.32 | 0.71 |
| Intervention | 6.23 | 6.50 | 0.27 | 6.23 | 6.70 | 0.47 |
| Incremental | 0.46 | 0.34 | -0.12 | 0.62 | 0.38 | -0.24 |
| *QALYs* |  |  |  |  |  |  |
| Standard of care | 2.98 | 2.94 | -0.04 | 3.68 | 4.08 | 0.40 |
| Intervention | 3.49 | 3.29 | -0.20 | 4.32 | 4.49 | 0.17 |
| Incremental | 0.51 | 0.35 | -0.16 | 0.64 | 0.41 | -0.23 |
| *Total costs* |  |  |  |  |  |  |
| Standard of care | \$670,000 | \$484,963 | \$-185,037 | \$390,153 | \$438,806 | \$48,653 |
| Intervention | \$790,000 | \$580,334 | \$-209,666 | \$382,702 | \$435,516 | \$52,814 |
| Incremental | \$120,000 | \$95,370 | \$-24,630 | \$-7,451 | \$-3,290 | \$4,161 |
| *Incremental cost-effectiveness ratio* | \$236,000 | \$272,131 | \$36,131 | Not applicable | Not applicable | Not applicable |

Abbreviations: QALY, quality-adjusted life years.
\* Lecanemab drug costs were not included in the original model and the IPECAD cross-validation.

### USE CASE 2: CROSS-VALIDATE EXISTING AD-ACE MODEL

Where possible, features of the AD-ACE model were implemented except its structure and treatment effect waning due to its integration with structure. The reason was the fundamental difference in structure with AD-ACE a patient-level microsimulation on continuous disease progression domains cognition, function, behavior and biomarkers and IPECAD a cohort state transition on discrete disease progression composite domain of cognition and function. See **supplemental material Table S1** and its table notes for a list of all input parameters and short description.

Compared to AD-ACE the IPECAD model outcomes showed higher life expectancy (+0.71 year), higher QALYs (+0.40) and higher costs (+$49,000) in the standard of care strategy. Incremental life years, QALY gains and cost savings were smaller (36-56% smaller) (see **Table 1**).

### USE CASE 3: ADDITIONAL UNCERTAINTY SCENARIOS

Alternative scenarios were tested in univariate sensitivity analyses. First, the lecanemab trial primary efficacy endpoint of ‘relative difference in CDR-SB change from baseline’ and non-reported endpoint of ‘time shift in CDR-SB change from baseline’ was used as alternative to the ‘hazard ratio for progression to the next stage of dementia’. Second, an alternative source for natural progression from MCI to dementia was obtained from Vos et al. (39), and for natural progression between dementia states and death was obtained from Wimo et al. (23). Third, alternative extrapolating beyond the trial follow-up period were addressed in terms of ad-hoc combinations of treatment discontinuation (i.e., stopping rules) and treatment effect waning rates (see **supplemental Figure S1**). Fourth, an alternative method of a shorter cycle length of 1/24 month was employed.

Results of these additional uncertainty scenarios are presented in **Table 2**. As a result of extrapolation beyond evidence from the trial follow-up period more than half of the costs fell in the first 2 years while the majority of the benefits (QALY gain and care savings) were achieved after 2 years. Cost-effectiveness was lower (i.e., less net health benefit) in scenarios reflecting slower progression and treatment effect waning during treatment, and was higher (i.e., more net health benefit) in scenarios reflecting stopping treatment while assuming a treatment effect after stopping. We note the lack of evidence for stopping and waning assumptions related to the latter results.

**Table 2:**
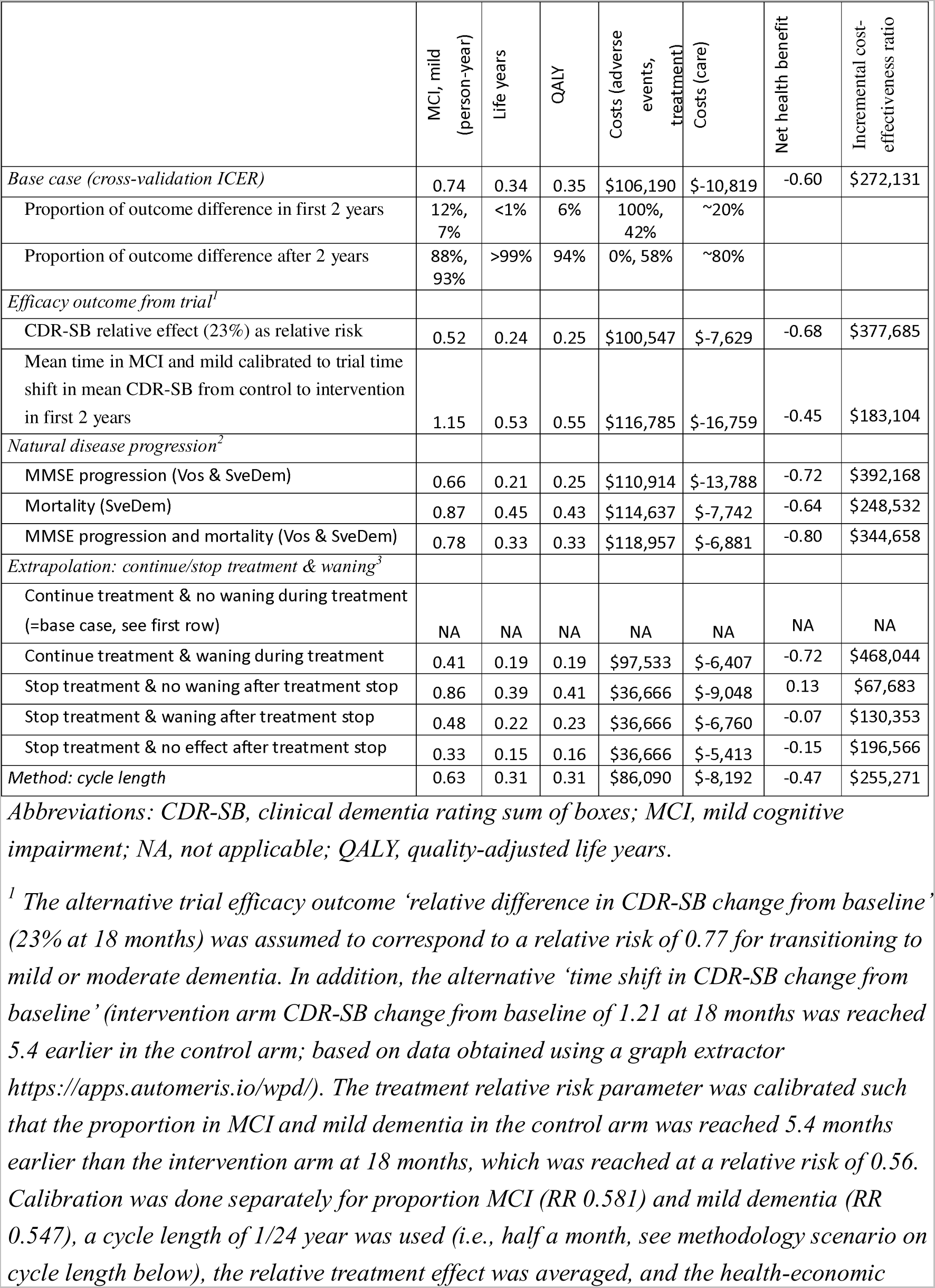
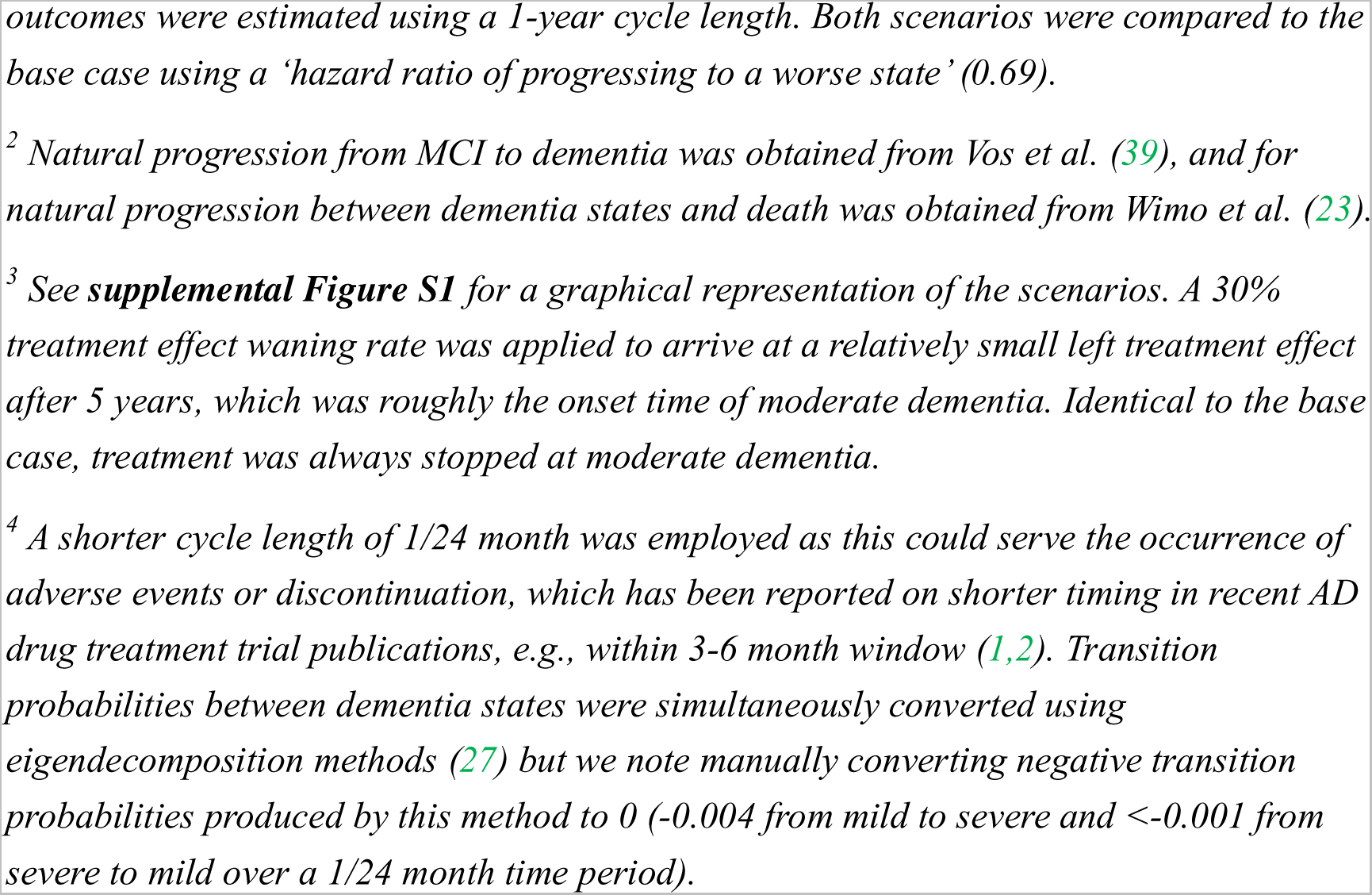
Difference (intervention minus standard of care strategy) in model outcomes of sensitivity analyses (person-years undiscounted; QALY, costs and net health benefit discounted in US dollar). See supplemental material Table Sx for details on the scenarios.

|  | MCI, mild<br>(person-year) | Life years | QALY | Costs (adverse<br>events,<br>treatment) | Costs (care) | Net health benefit | Incremental cost-effectiveness ratio |
| --- | --- | --- | --- | --- | --- | --- | --- |
| *Base case (cross-validation ICER)* | 0.74 | 0.34 | 0.35 | \$106,190 | \$-10,819 | -0.60 | \$272,131 |
| Proportion of outcome difference in first 2 years | 12%,<br>7% | <1% | 6% | 100%,<br>42% | ~20% |  |  |
| Proportion of outcome difference after 2 years | 88%,<br>93% | >99% | 94% | 0%, 58% | ~80% |  |  |
| *Efficacy outcome from trial<sup>1</sup>* |  |  |  |  |  |  |  |
| CDR-SB relative effect (23%) as relative risk | 0.52 | 0.24 | 0.25 | \$100,547 | \$-7,629 | -0.68 | \$377,685 |
| Mean time in MCI and mild calibrated to trial time shift in mean CDR-SB from control to intervention in first 2 years | 1.15 | 0.53 | 0.55 | \$116,785 | \$-16,759 | -0.45 | \$183,104 |
| *Natural disease progression<sup>2</sup>* |  |  |  |  |  |  |  |
| MMSE progression (Vos & SveDem) | 0.66 | 0.21 | 0.25 | \$110,914 | \$-13,788 | -0.72 | \$392,168 |
| Mortality (SveDem) | 0.87 | 0.45 | 0.43 | \$114,637 | \$-7,742 | -0.64 | \$248,532 |
| MMSE progression and mortality (Vos & SveDem) | 0.78 | 0.33 | 0.33 | \$118,957 | \$-6,881 | -0.80 | \$344,658 |
| *Extrapolation: continue/stop treatment & waning<sup>3</sup>* |  |  |  |  |  |  |  |
| Continue treatment & no waning during treatment (=base case, see first row) | NA | NA | NA | NA | NA | NA | NA |
| Continue treatment & waning during treatment | 0.41 | 0.19 | 0.19 | \$97,533 | \$-6,407 | -0.72 | \$468,044 |
| Stop treatment & no waning after treatment stop | 0.86 | 0.39 | 0.41 | \$36,666 | \$-9,048 | 0.13 | \$67,683 |
| Stop treatment & waning after treatment stop | 0.48 | 0.22 | 0.23 | \$36,666 | \$-6,760 | -0.07 | \$130,353 |
| Stop treatment & no effect after treatment stop | 0.33 | 0.15 | 0.16 | \$36,666 | \$-5,413 | -0.15 | \$196,566 |
| *Method: cycle length* | 0.63 | 0.31 | 0.31 | \$86,090 | \$-8,192 | -0.47 | \$255,271 |

*Abbreviations: CDR-SB, clinical dementia rating sum of boxes; MCI, mild cognitive impairment; NA, not applicable; QALY, quality-adjusted life years.*
<sup>1</sup> The alternative trial efficacy outcome ‘relative difference in CDR-SB change from baseline’ (23% at 18 months) was assumed to correspond to a relative risk of 0.77 for transitioning to mild or moderate dementia. In addition, the alternative ‘time shift in CDR-SB change from baseline’ (intervention arm CDR-SB change from baseline of 1.21 at 18 months was reached 5.4 earlier in the control arm; based on data obtained using a graph extractor https://apps.automeris.io/wpd/). The treatment relative risk parameter was calibrated such that the proportion in MCI and mild dementia in the control arm was reached 5.4 months earlier than the intervention arm at 18 months, which was reached at a relative risk of 0.56. Calibration was done separately for proportion MCI (RR 0.581) and mild dementia (RR 0.547), a cycle length of 1/24 year was used (i.e., half a month, see methodology scenario on cycle length below), the relative treatment effect was averaged, and the health-economic
outcomes were estimated using a 1-year cycle length. Both scenarios were compared to the base case using a ‘hazard ratio of progressing to a worse state’ (0.69).
<sup>2</sup> Natural progression from MCI to dementia was obtained from Vos et al. (39), and for natural progression between dementia states and death was obtained from Wimo et al. (23).
<sup>3</sup> See **supplemental Figure S1** for a graphical representation of the scenarios. A 30% treatment effect waning rate was applied to arrive at a relatively small left treatment effect after 5 years, which was roughly the onset time of moderate dementia. Identical to the base case, treatment was always stopped at moderate dementia.
<sup>4</sup> A shorter cycle length of 1/24 month was employed as this could serve the occurrence of adverse events or discontinuation, which has been reported on shorter timing in recent AD drug treatment trial publications, e.g., within 3-6 month window (1,2). Transition probabilities between dementia states were simultaneously converted using eigendecomposition methods (27) but we note manually converting negative transition probabilities produced by this method to 0 (-0.004 from mild to severe and <-0.001 from severe to mild over a 1/24 month time period).

## DISCUSSION

We present a new IPECAD open-source model framework (version 2) for the health-economic evaluation of early AD treatment. We demonstrated its ability for cross-validation of other published analyses as well as additional uncertainty analyses. Health-economic outcomes seem related to model type and assumptions on treatment stopping rules and effect waning.

Differences with the reported ICER model outcomes could be explained by some features (e.g., start in institution care setting or age-specific utility and costs) not implemented in our IPECAD model. There may also have been ICER model features we interpreted or programmed differently. We note the SveDem model was closely replicated and post-hoc we relatively closely replicated another disease progression model by Herring et al. (14) (see **supplemental Table S3**). However, these two replications relied on support from the original developers. We think the limited replicability creates an opportunity for open-source modeling to improve transparency on model details.

Differences with the reported AD-ACE model outcomes could result from the assumption in AD-ACE that eventually patients would be in a similar state as if they had not been treated, which implies compression rather than postponing of the time spent in severe dementia. This would seemingly imply similar mortality between intervention and control, and thus fewer QALYs gained related to extending life. However, the AD-ACE model reported a larger QALY gain as compared to our IPECAD cross-validation. There may also have been AD-ACE model features we interpreted or programmed differently.

The additional uncertainty analyses showed that health-economic outcomes were sensitivity to assumptions on treatment stopping rules and treatment effect waning when extrapolating from the lecanemab trial 18-month follow-up period. We note calibrating the model to the time shift on the primary trial outcome required adjusting the model cycle length, which was also subject to uncertainty. In addition, the time shift scenario relied on the assumption that all aspects of the disease are shifted in time (i.e., a time shift in change from baseline on a continuous outcome translates to the same time shift in proportion in disease severity state).

The uncertainty scenarios confirm previous model comparison studies showing health-economic outcomes are sensitive to the choice of efficacy outcome from the trial and choice of natural disease history source (16, 17). Among recently reviewed studies (5) none addressed cycle time, and few addressed treatment discontinuation and treatment effect waning in scenario and sensitivity analyses. Whittington et al. (40) showed a lower incremental cost-effectiveness ratio with aducanumab treatment stopping at an earlier severity state and diminished outcomes with stronger treatment effect waning assumptions. AD-ACE model applications (22, 34, 41) showed higher lecanemab treatment value with higher discontinuation rates, with maintained reduced amyloid level after stopping treatment and with lower dosing frequency while assuming no treatment effect waning, and lower lecanemab treatment value with shorter maximum treatment duration. Kongpakwattana et al. (42) showed a higher cost-effectiveness ratio with earlier stopping donepezil at severe dementia. Ross et al. (15) showed a higher cost-effectiveness ratio with later discontinuation at severe instead of moderate dementia. An additional identified study (43) showed a lower ICER with a shorter maximum treatment duration, a lower ICER when assuming sustained effect after discontinuation due to amyloid clearance and a lower ICER when assuming treatment effects sustained longer. Another additional study (44) showed a higher ICER when assuming treatment effect waning. For the majority these results overlap to our finding of higher net benefit for higher discontinuation and optimistic treatment waning assumptions.

Recent cross-comparison studies in AD cross-validated models by comparing them after implementing a common benchmark scenario (16, 17). Although most previous modeling studies compared model outcomes to other studies (19), we identified only one example cross-validating a model by partly implementing another model’s scenario (14) in terms of starting age. We confirm the observation that differences between models are difficult to explain. Therefore, we advocate standardized reporting of undiscounted non-half-cycle-adjusted proportions in states over time to improve comparability of model outcomes to understand their differences (see **supplemental Table S2**) and sharing model outcomes using open science principles (for example on IPECAD repository https://osf.io/jv85a).

### IMPLICATIONS FOR REIMBURSEMENT AGENCIES AND MODELERS

We think the new IPECAD open-source model framework can support transparency and credibility of new cost-effectiveness assessments for new early AD treatments. First, the model can be copied, adjusted or further developed with little effort and (staff or time) resources. Second, the model can cross-validate a model submitted by industry to increase the scope and rigor. Third, the model can address additional (uncertainty or subgroup) analyses, for example addressing uncertainties that have received little attention in previous research. We note addressing parameter uncertainty fell outside our scope but can be relatively easily implemented. Fourth, the model can be used for other (educational) purposes without any risk of confidential information being compromised.

In addition, our study results showed uncertainty related to assumptions on treatment stopping rules and effect waning. Because a detailed elaboration of these aspects falls outside the scope of our study, we recommend addressing in the assessment of lecanemab, donanemab and future AD treatments.

### LIMITATIONS

Our study is subject to several limitations. We did not engage with patients and other stakeholders for the conceptualization of our model. However, another study has provided a rationale for the choice of a state-transition model type with similar disease states (30).

The model lacks reflecting diagnostic infrastructure to identify persons with abnormal amyloid eligible for treatment (45, 46), although this could be reflected by a number needed to test to identify a person eligible for treatment and corresponding test costs. Also, the model does not estimate the potential budget impact of new AD treatments, which likely is important to decision makers (6, 47). Also, the model does not reflect treatment effectiveness in specific subgroups such as APOE4, which could be associated to treatment effectiveness, adverse events and test costs.

The open-source nature of the model does not imply the validity of the model or any of its applications and does not imply it is error-free. Valid use implies adhering to scientific integrity standards for example in terms of transparency in selection of model inputs and assumptions (as compared to incorrect or ‘off-the-shelf’ application) (9). For example, careful consideration of implementing trial efficacy outcomes into the model as well as the choice for natural progression is important (17). Nevertheless, the open-source nature has the potential to reduce the presence of technical errors and facilitate incremental improvements (9), as well as to reduce the room for interpretation making it practically fully replicable.

Our model structure is of relatively simple nature, missing features for example to reflect treatment switching, domain-specific effects, alternative assumptions for mortality, efficiently address heterogeneity, treatment in institutional setting and time-dependent transitions between disease states. Also, we did not cross-validate the ICER state transition model with a microsimulation-type model. We note our previous IPECAD open-source model framework microsimulation version (version 1.2 available on www.ipecad.org) allows some of these features (48). Alternatively, the open-source nature of our model allows adding new features such as time-specific transitions (26) by skilled programmers.

### CONCLUSION

A new IPECAD open-source model framework (version 2) was developed for the health-economic evaluation of early AD treatment. We demonstrated its ability for researchers and decision-makers to cross-validate other (HTA submission) models and perform additional uncertainty analyses, setting an example for open science in AD decision modeling and supporting important reimbursement decisions.

## AUTHOR CONTRIBUTIONS

Concept and design: RH, AW

Development of the model: RH

Applications of the model: RH, WLH, SG, LJ

Analysis and interpretation of data: RH, WLH, SG

Drafting the manuscript: RH

Critical revision: ALL

## Data Availability

All data produced are available online at https://github.com/ronhandels/IPECAD

## ACKNOWLEDGMENT

We wish to acknowledge master students Linh Nguyen (BSc) and Daphne Silvertand (BSc) for their support in the development of the model.

## CONFLICT OF INTEREST

RH received outside this study research grants from JPND, ZonMW, IMI, H2020 (paid to institution); received outside this study consulting fees in the past 3 years from Lilly Nederland (2023), iMTA (2023), and Biogen (2021) (paid to institution); is member of IPECAD and member of ISPOR special interest group open-source models (un-paid).

WLH is a full-time employee of RTI Health Solutions, an independent nonprofit research organization. His compensation is unconnected to the studies on which he works. RTI Health Solutions did not receive funding for the current study. Outside of this study, RTI Health Solutions receives funding pursuant to research contracts with pharmaceutical and biotechnology companies. WLH is affiliated to research at the Karolinska Institutet and receives no compensation as part of this affiliation.

SG: has no conflict of interest to declare.

AW reports License holder of RUD-instrument (part) and Alzheimer Disease International MSAP scientific committee.

AS has no conflict of interest to declare.

LJ received outside of this study consulting fees from H. Lundbeck A/S, Novo Nordisk AS, Eli Lilly Inc and license fees for the Resource Utilization in Dementia (RUD) instrument.

BW received license fees for the Resource Utilization in Dementia (RUD) instrument.

## SUPPLEMENTAL MATERIAL

Supplemental material related to manuscript entitled “New IPECAD open-source model framework for the health technology assessment of early Alzheimer’s disease treatment: development and use cases”.

**Table S1:**
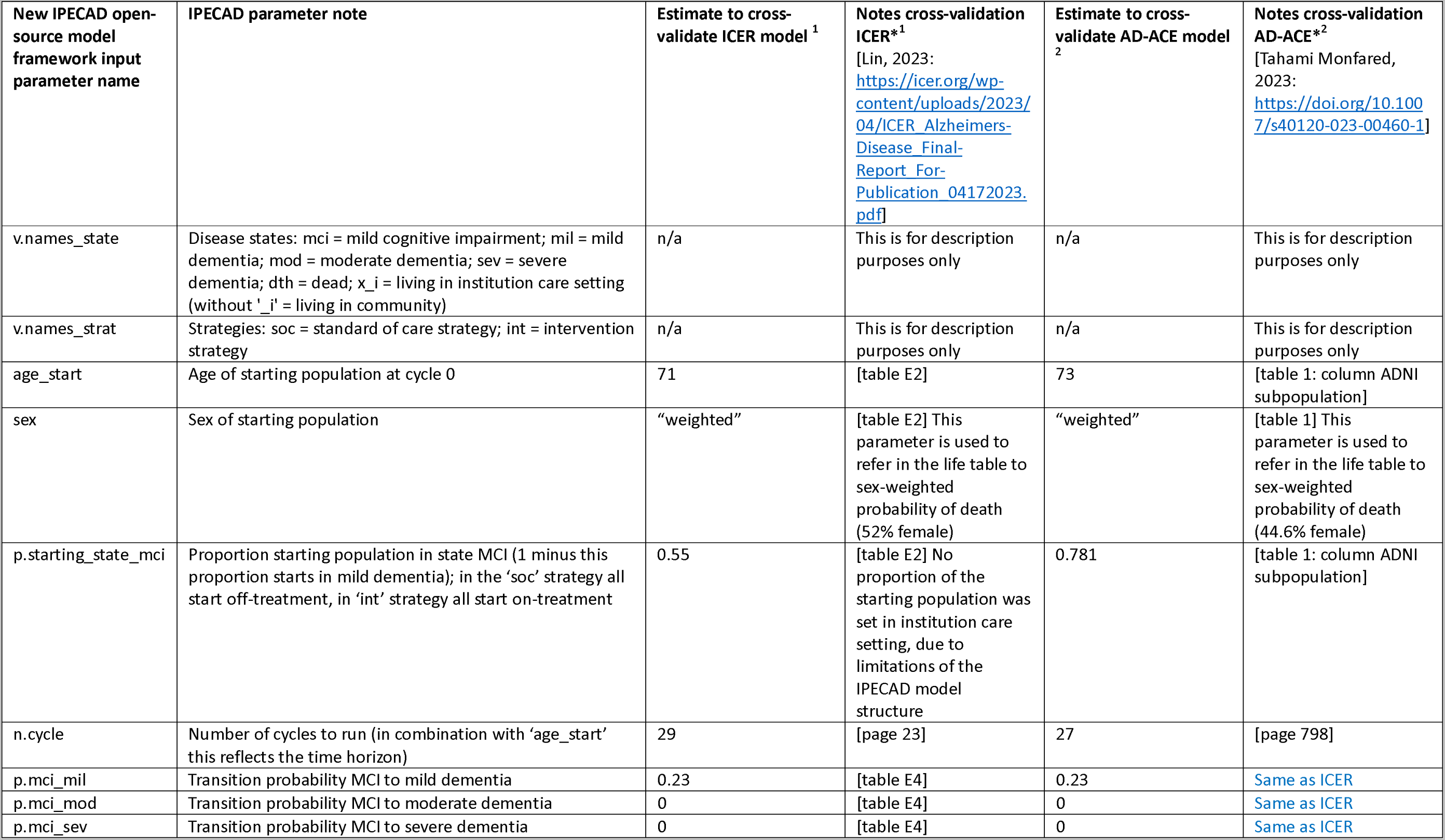
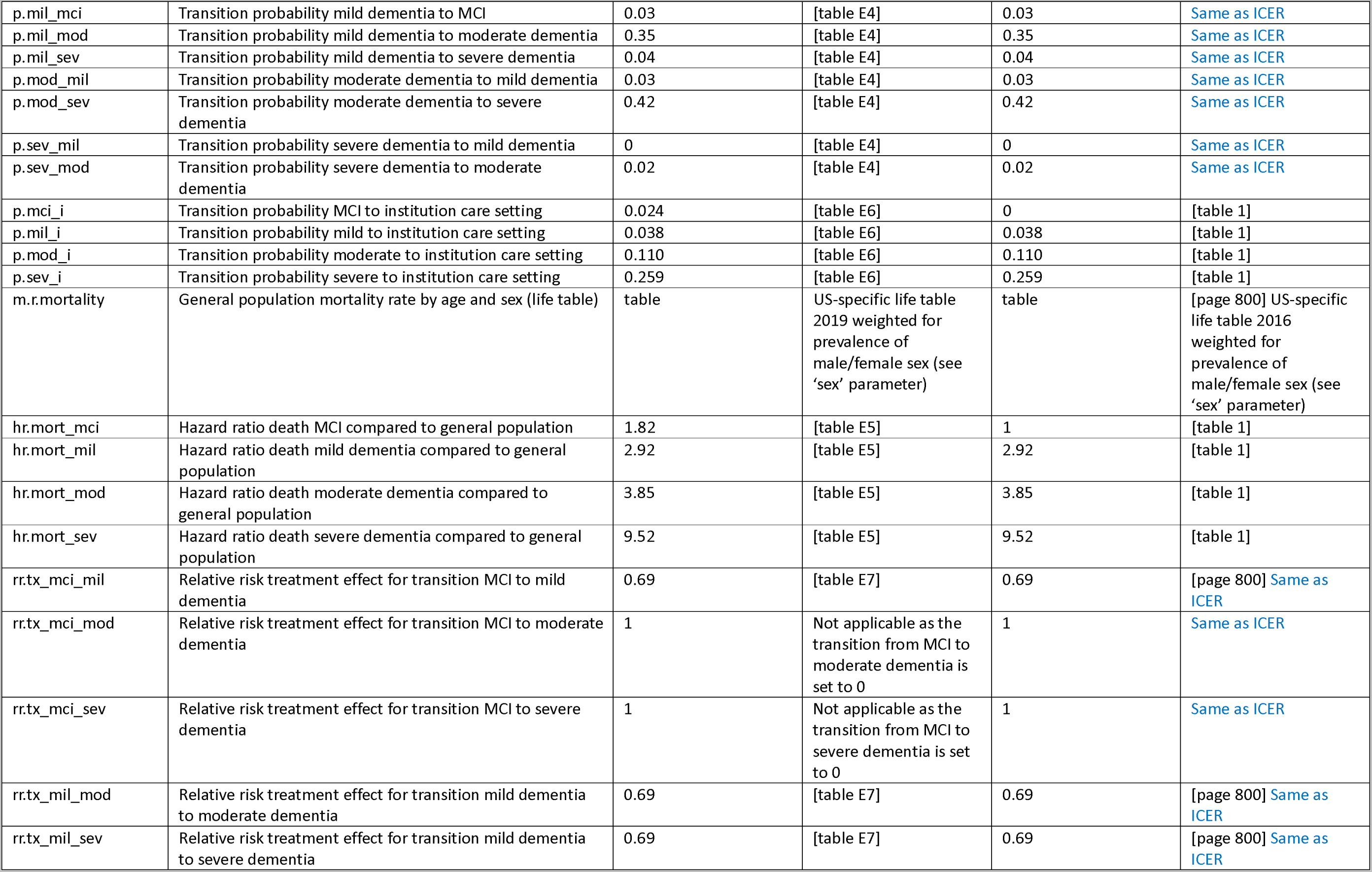
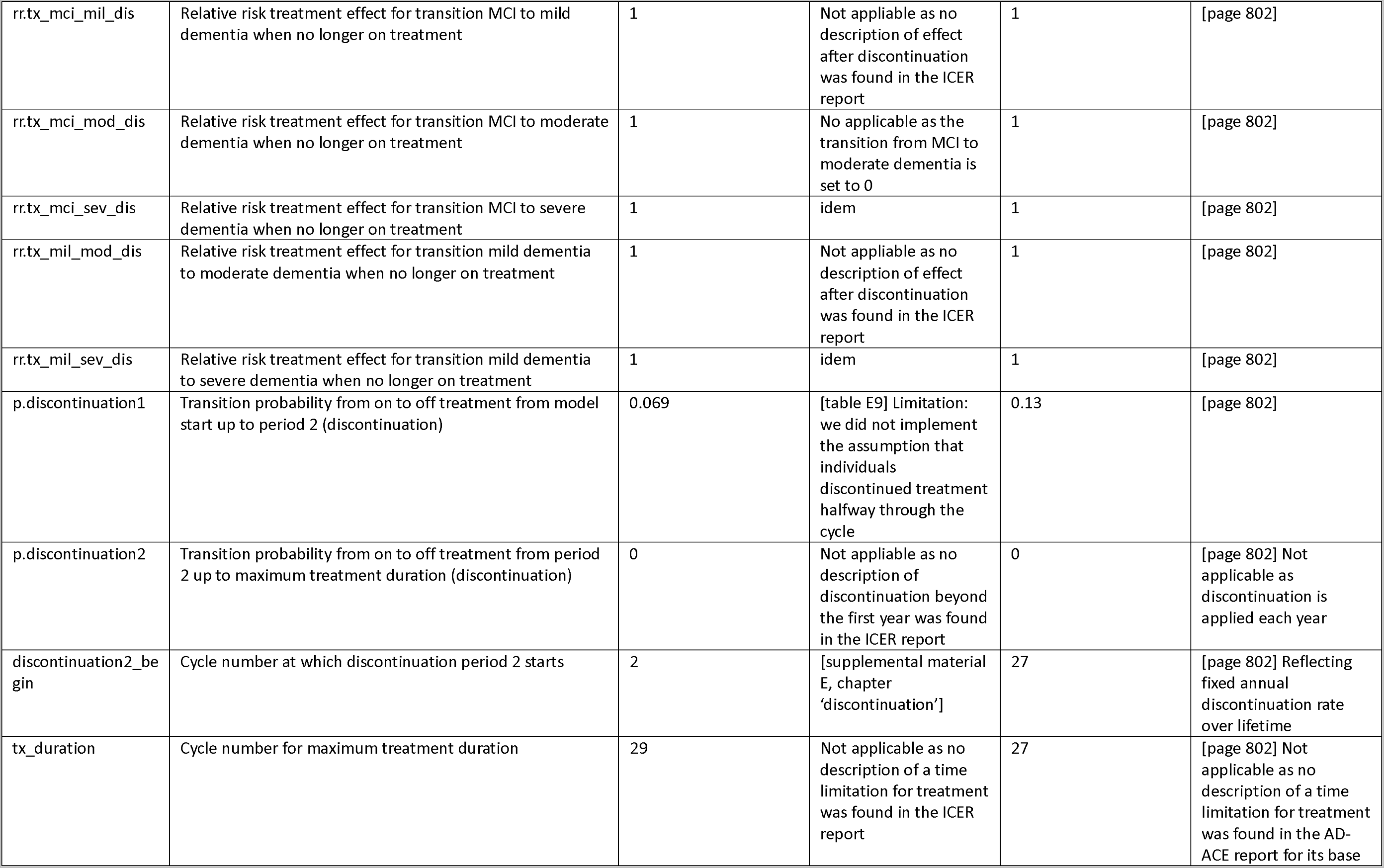
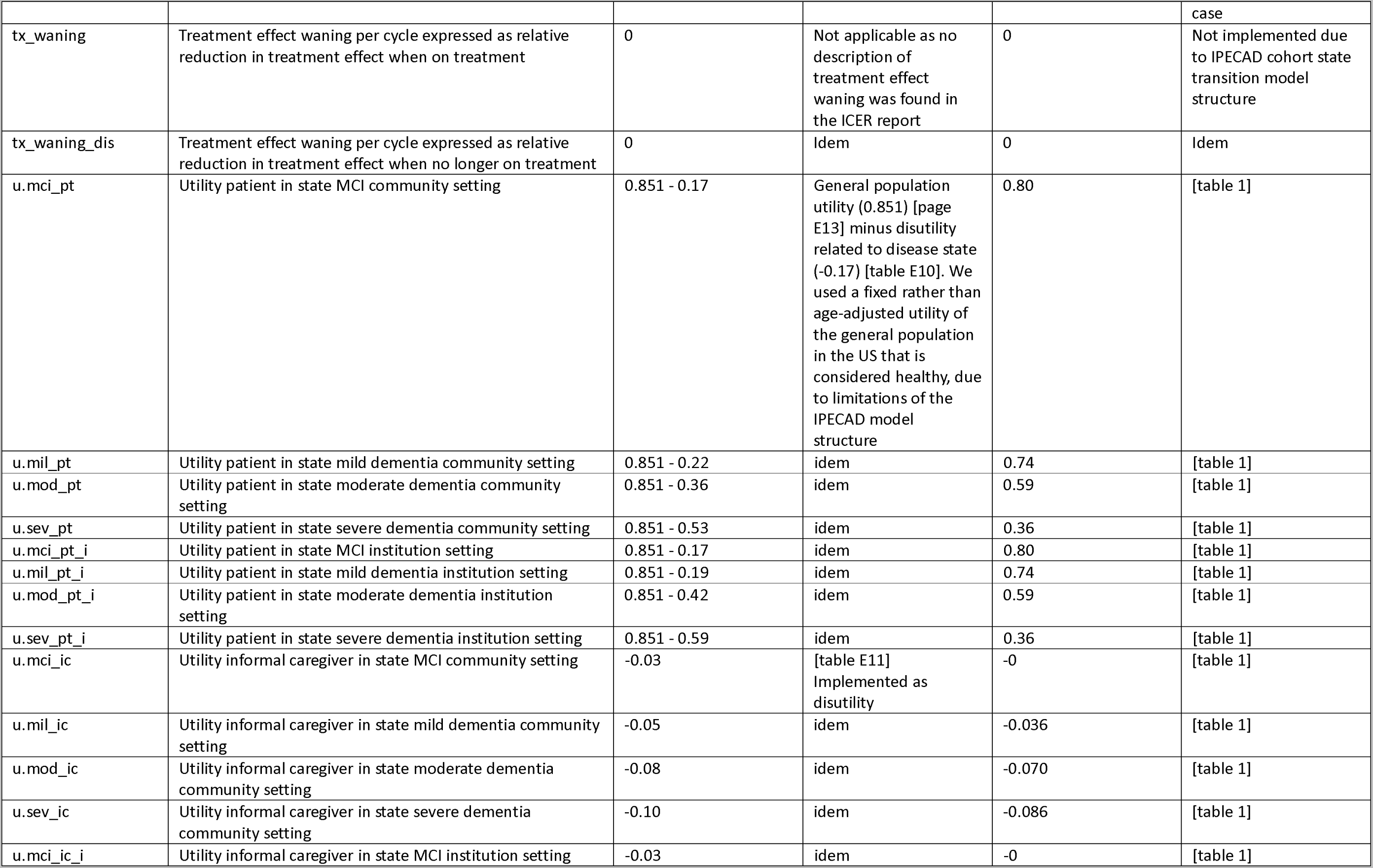
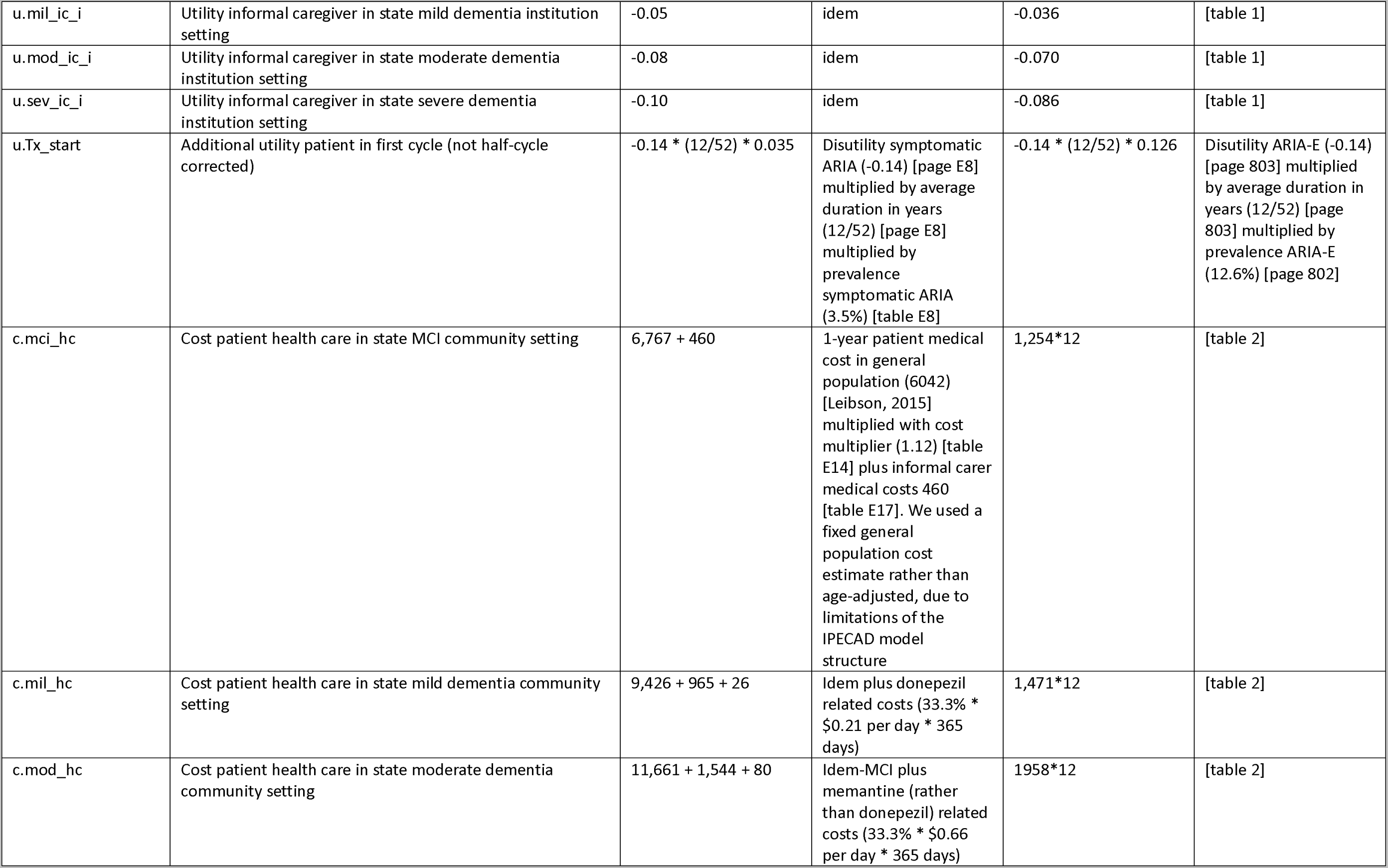
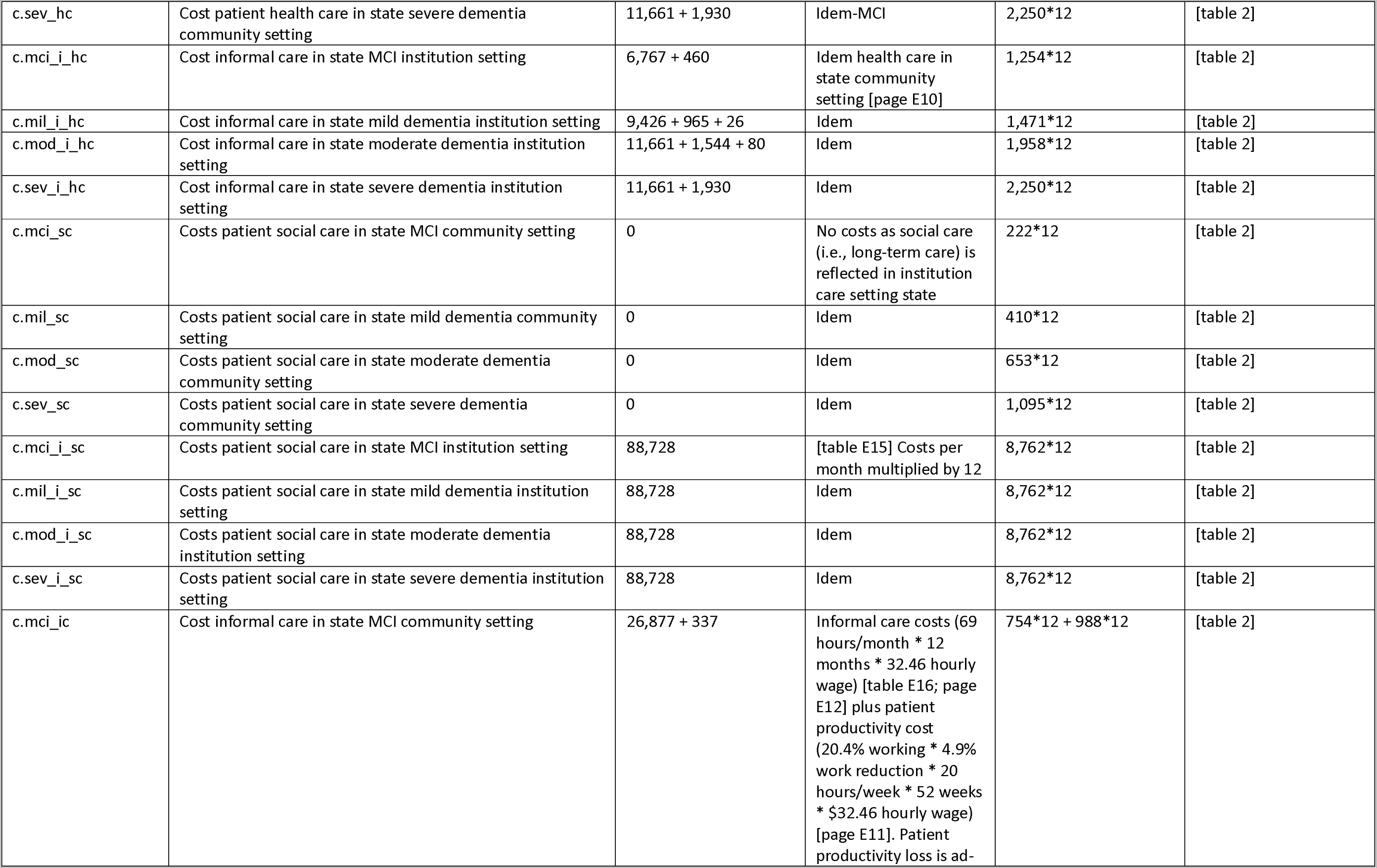
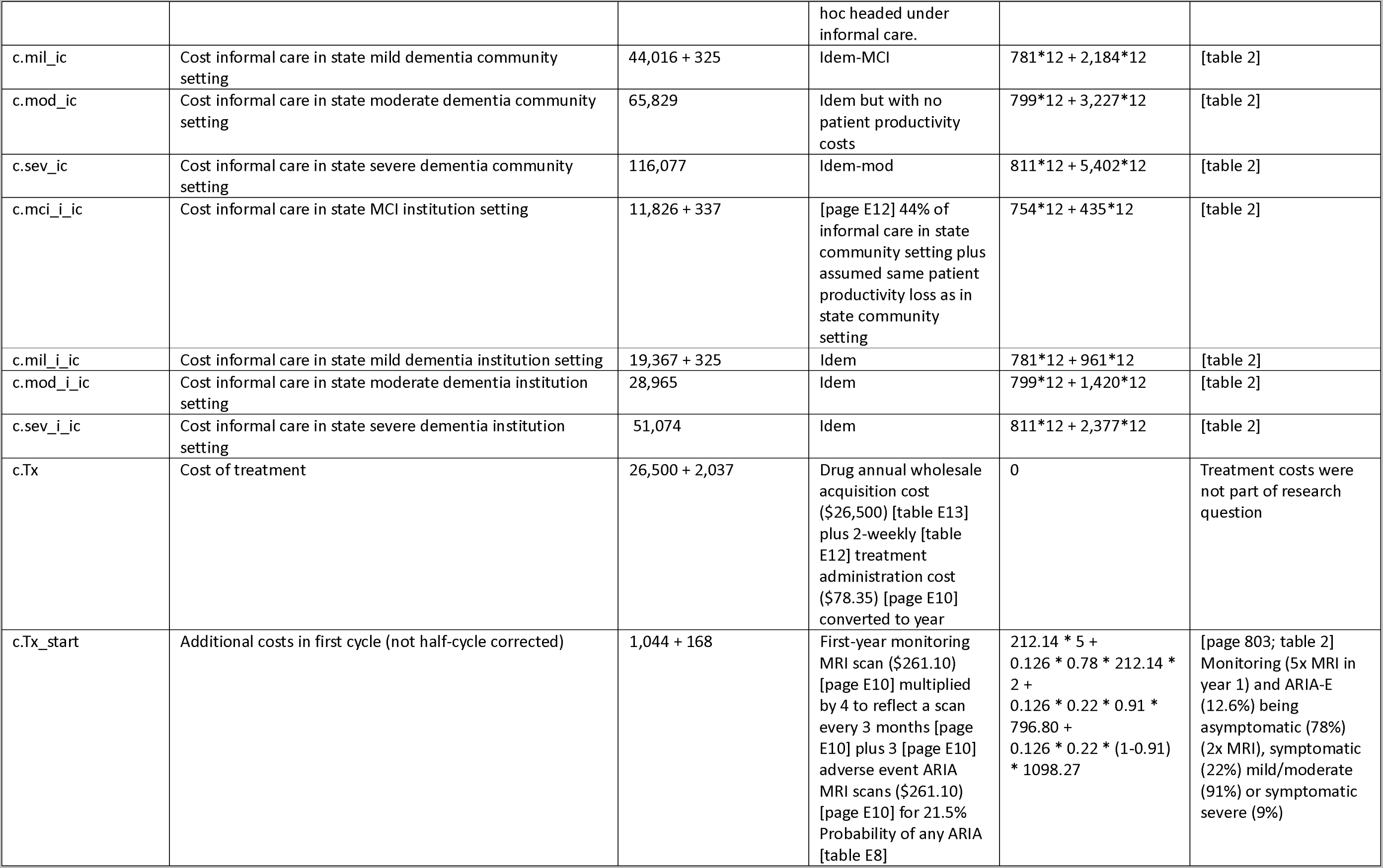
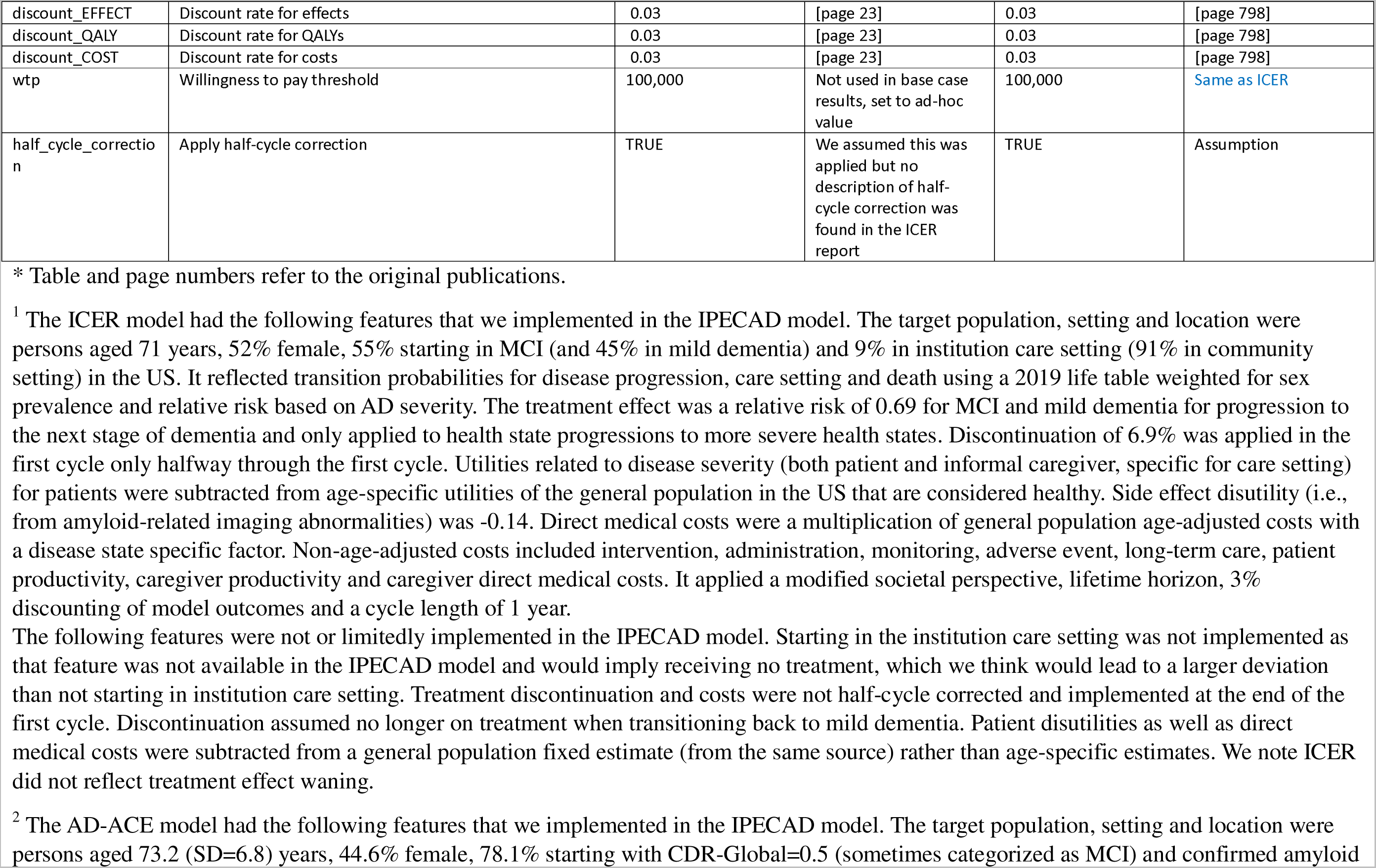
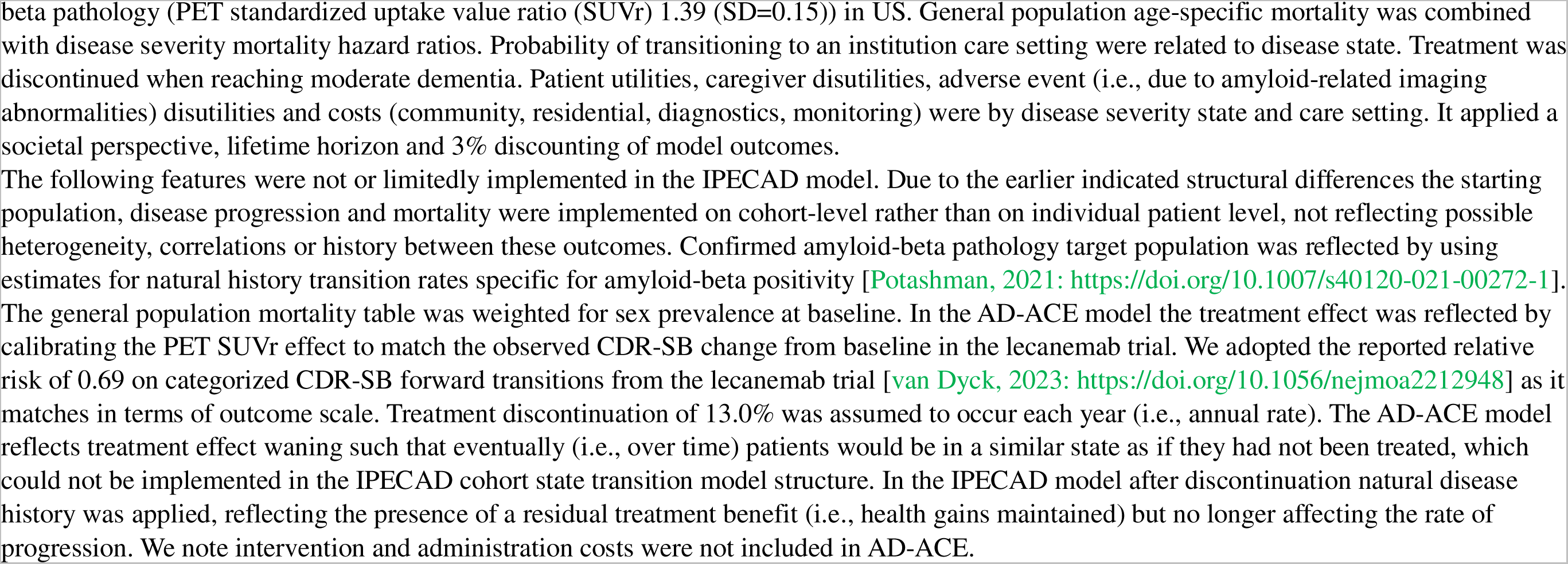
Summary of the input parameters of the new IPECAD open-source model, the estimate to reflect the use case and related comments. Inputs are per cycle unless specified otherwise, with cycle length of 1 year. Page and table numbers refer to citation in column header.

| New IPECAD open-source model framework input parameter name | IPECAD parameter note | Estimate to cross-validate ICER model <sup>1</sup> | Notes cross-validation ICER*<sup>1</sup><br>[Lin, 2023: https://icer.org/wp-content/uploads/2023/04/ICER_Alzheimers-Disease_Final-Report_For-Publication_04172023.pdf] | Estimate to cross-validate AD-ACE model <sup>2</sup> | Notes cross-validation AD-ACE*<sup>2</sup><br>[Tahami Monfared, 2023: https://doi.org/10.1007/s40120-023-00460-1] |
| --- | --- | --- | --- | --- | --- |
| v.names_state | Disease states: mci = mild cognitive impairment; mil = mild dementia; mod = moderate dementia; sev = severe dementia; dth = dead; x_i = living in institution care setting (without 'i' = living in community) | n/a | This is for description purposes only | n/a | This is for description purposes only |
| v.names_strat | Strategies: soc = standard of care strategy; int = intervention strategy | n/a | This is for description purposes only | n/a | This is for description purposes only |
| age_start | Age of starting population at cycle 0 | 71 | [table E2] | 73 | [table 1: column ADNI subpopulation] |
| sex | Sex of starting population | “weighted” | [table E2] This parameter is used to refer in the life table to sex-weighted probability of death (52% female) | “weighted” | [table 1] This parameter is used to refer in the life table to sex-weighted probability of death (44.6% female) |
| p.starting_state_mci | Proportion starting population in state MCI (1 minus this proportion starts in mild dementia); in the ‘soc’ strategy all start off-treatment, in ‘int’ strategy all start on-treatment | 0.55 | [table E2] No proportion of the starting population was set in institution care setting, due to limitations of the IPECAD model structure | 0.781 | [table 1: column ADNI subpopulation] |
| n.cycle | Number of cycles to run (in combination with ‘age_start’ this reflects the time horizon) | 29 | [page 23] | 27 | [page 798] |
| p.mci_mil | Transition probability MCI to mild dementia | 0.23 | [table E4] | 0.23 | Same as ICER |
| p.mci_mod | Transition probability MCI to moderate dementia | 0 | [table E4] | 0 | Same as ICER |
| p.mci_sev | Transition probability MCI to severe dementia | 0 | [table E4] | 0 | Same as ICER |
| p.mil_mci | Transition probability mild dementia to MCI | 0.03 | [table E4] | 0.03 | Same as ICER |
| p.mil_mod | Transition probability mild dementia to moderate dementia | 0.35 | [table E4] | 0.35 | Same as ICER |
| p.mil_sev | Transition probability mild dementia to severe dementia | 0.04 | [table E4] | 0.04 | Same as ICER |
| p.mod_mil | Transition probability moderate dementia to mild dementia | 0.03 | [table E4] | 0.03 | Same as ICER |
| p.mod_sev | Transition probability moderate dementia to severe dementia | 0.42 | [table E4] | 0.42 | Same as ICER |
| p.sev_mil | Transition probability severe dementia to mild dementia | 0 | [table E4] | 0 | Same as ICER |
| p.sev_mod | Transition probability severe dementia to moderate dementia | 0.02 | [table E4] | 0.02 | Same as ICER |
| p.mci_i | Transition probability MCI to institution care setting | 0.024 | [table E6] | 0 | [table 1] |
| p.mil_i | Transition probability mild to institution care setting | 0.038 | [table E6] | 0.038 | [table 1] |
| p.mod_i | Transition probability moderate to institution care setting | 0.110 | [table E6] | 0.110 | [table 1] |
| p.sev_i | Transition probability severe to institution care setting | 0.259 | [table E6] | 0.259 | [table 1] |
| m.r.mortality | General population mortality rate by age and sex (life table) | table | US-specific life table 2019 weighted for prevalence of male/female sex (see 'sex' parameter) | table | [page 800] US-specific life table 2016 weighted for prevalence of male/female sex (see 'sex' parameter) |
| hr.mort_mci | Hazard ratio death MCI compared to general population | 1.82 | [table E5] | 1 | [table 1] |
| hr.mort_mil | Hazard ratio death mild dementia compared to general population | 2.92 | [table E5] | 2.92 | [table 1] |
| hr.mort_mod | Hazard ratio death moderate dementia compared to general population | 3.85 | [table E5] | 3.85 | [table 1] |
| hr.mort_sev | Hazard ratio death severe dementia compared to general population | 9.52 | [table E5] | 9.52 | [table 1] |
| rr.tx_mci_mil | Relative risk treatment effect for transition MCI to mild dementia | 0.69 | [table E7] | 0.69 | [page 800] Same as ICER |
| rr.tx_mci_mod | Relative risk treatment effect for transition MCI to moderate dementia | 1 | Not applicable as the transition from MCI to moderate dementia is set to 0 | 1 | Same as ICER |
| rr.tx_mci_sev | Relative risk treatment effect for transition MCI to severe dementia | 1 | Not applicable as the transition from MCI to severe dementia is set to 0 | 1 | Same as ICER |
| rr.tx_mil_mod | Relative risk treatment effect for transition mild dementia to moderate dementia | 0.69 | [table E7] | 0.69 | [page 800] Same as ICER |
| rr.tx_mil_sev | Relative risk treatment effect for transition mild dementia to severe dementia | 0.69 | [table E7] | 0.69 | [page 800] Same as ICER |
| rr.tx_mci_mil_dis | Relative risk treatment effect for transition MCI to mild dementia when no longer on treatment | 1 | Not applicable as no description of effect after discontinuation was found in the ICER report | 1 | [page 802] |
| rr.tx_mci_mod_dis | Relative risk treatment effect for transition MCI to moderate dementia when no longer on treatment | 1 | No applicable as the transition from MCI to moderate dementia is set to 0 | 1 | [page 802] |
| rr.tx_mci_sev_dis | Relative risk treatment effect for transition MCI to severe dementia when no longer on treatment | 1 | idem | 1 | [page 802] |
| rr.tx_mil_mod_dis | Relative risk treatment effect for transition mild dementia to moderate dementia when no longer on treatment | 1 | Not applicable as no description of effect after discontinuation was found in the ICER report | 1 | [page 802] |
| rr.tx_mil_sev_dis | Relative risk treatment effect for transition mild dementia to severe dementia when no longer on treatment | 1 | idem | 1 | [page 802] |
| p.discontinuation1 | Transition probability from on to off treatment from model start up to period 2 (discontinuation) | 0.069 | [table E9] Limitation: we did not implement the assumption that individuals discontinued treatment halfway through the cycle | 0.13 | [page 802] |
| p.discontinuation2 | Transition probability from on to off treatment from period 2 up to maximum treatment duration (discontinuation) | 0 | Not applicable as no description of discontinuation beyond the first year was found in the ICER report | 0 | [page 802] Not applicable as discontinuation is applied each year |
| discontinuation2_begin | Cycle number at which discontinuation period 2 starts | 2 | [supplemental material E, chapter 'discontinuation'] | 27 | [page 802] Reflecting fixed annual discontinuation rate over lifetime |
| tx_duration | Cycle number for maximum treatment duration | 29 | Not applicable as no description of a time limitation for treatment was found in the ICER report | 27 | [page 802] Not applicable as no description of a time limitation for treatment was found in the AD-ACE report for its base case |
| tx_waning | Treatment effect waning per cycle expressed as relative reduction in treatment effect when on treatment | 0 | Not applicable as no description of treatment effect waning was found in the ICER report | 0 | Not implemented due to IPECAD cohort state transition model structure |
| tx_waning_dis | Treatment effect waning per cycle expressed as relative reduction in treatment effect when no longer on treatment | 0 | Idem | 0 | Idem |
| u.mci_pt | Utility patient in state MCI community setting | 0.851 - 0.17 | General population utility (0.851) [page E13] minus disutility related to disease state (-0.17) [table E10]. We used a fixed rather than age-adjusted utility of the general population in the US that is considered healthy, due to limitations of the IPECAD model structure | 0.80 | [table 1] |
| u.mil_pt | Utility patient in state mild dementia community setting | 0.851 - 0.22 | idem | 0.74 | [table 1] |
| u.mod_pt | Utility patient in state moderate dementia community setting | 0.851 - 0.36 | idem | 0.59 | [table 1] |
| u.sev_pt | Utility patient in state severe dementia community setting | 0.851 - 0.53 | idem | 0.36 | [table 1] |
| u.mci_pt_i | Utility patient in state MCI institution setting | 0.851 - 0.17 | idem | 0.80 | [table 1] |
| u.mil_pt_i | Utility patient in state mild dementia institution setting | 0.851 - 0.19 | idem | 0.74 | [table 1] |
| u.mod_pt_i | Utility patient in state moderate dementia institution setting | 0.851 - 0.42 | idem | 0.59 | [table 1] |
| u.sev_pt_i | Utility patient in state severe dementia institution setting | 0.851 - 0.59 | idem | 0.36 | [table 1] |
| u.mci_ic | Utility informal caregiver in state MCI community setting | -0.03 | [table E11]<br>Implemented as disutility | -0 | [table 1] |
| u.mil_ic | Utility informal caregiver in state mild dementia community setting | -0.05 | idem | -0.036 | [table 1] |
| u.mod_ic | Utility informal caregiver in state moderate dementia community setting | -0.08 | idem | -0.070 | [table 1] |
| u.sev_ic | Utility informal caregiver in state severe dementia community setting | -0.10 | idem | -0.086 | [table 1] |
| u.mci_ic_i | Utility informal caregiver in state MCI institution setting | -0.03 | idem | -0 | [table 1] |
| u.mil_ic_i | Utility informal caregiver in state mild dementia institution setting | -0.05 | idem | -0.036 | [table 1] |
| u.mod_ic_i | Utility informal caregiver in state moderate dementia institution setting | -0.08 | idem | -0.070 | [table 1] |
| u.sev_ic_i | Utility informal caregiver in state severe dementia institution setting | -0.10 | idem | -0.086 | [table 1] |
| u.Tx_start | Additional utility patient in first cycle (not half-cycle corrected) | $-0.14 * (12/52) * 0.035$ | Disutility symptomatic ARIA (-0.14) [page E8] multiplied by average duration in years (12/52) [page E8] multiplied by prevalence symptomatic ARIA (3.5%) [table E8] | $-0.14 * (12/52) * 0.126$ | Disutility ARIA-E (-0.14) [page 803] multiplied by average duration in years (12/52) [page 803] multiplied by prevalence ARIA-E (12.6%) [page 802] |
| c.mci_hc | Cost patient health care in state MCI community setting | 6,767 + 460 | 1-year patient medical cost in general population (6042) [Leibson, 2015] multiplied with cost multiplier (1.12) [table E14] plus informal carer medical costs 460 [table E17]. We used a fixed general population cost estimate rather than age-adjusted, due to limitations of the IPECAD model structure | 1,254*12 | [table 2] |
| c.mil_hc | Cost patient health care in state mild dementia community setting | 9,426 + 965 + 26 | Idem plus donepezil related costs (33.3% * \$0.21 per day * 365 days) | 1,471*12 | [table 2] |
| c.mod_hc | Cost patient health care in state moderate dementia community setting | 11,661 + 1,544 + 80 | Idem-MCI plus memantine (rather than donepezil) related costs (33.3% * \$0.66 per day * 365 days) | 1958*12 | [table 2] |
| c.sev_hc | Cost patient health care in state severe dementia community setting | 11,661 + 1,930 | Idem-MCI | 2,250*12 | [table 2] |
| c.mci_i_hc | Cost informal care in state MCI institution setting | 6,767 + 460 | Idem health care in state community setting [page E10] | 1,254*12 | [table 2] |
| c.mil_i_hc | Cost informal care in state mild dementia institution setting | 9,426 + 965 + 26 | Idem | 1,471*12 | [table 2] |
| c.mod_i_hc | Cost informal care in state moderate dementia institution setting | 11,661 + 1,544 + 80 | Idem | 1,958*12 | [table 2] |
| c.sev_i_hc | Cost informal care in state severe dementia institution setting | 11,661 + 1,930 | Idem | 2,250*12 | [table 2] |
| c.mci_sc | Costs patient social care in state MCI community setting | 0 | No costs as social care (i.e., long-term care) is reflected in institution care setting state | 222*12 | [table 2] |
| c.mil_sc | Costs patient social care in state mild dementia community setting | 0 | Idem | 410*12 | [table 2] |
| c.mod_sc | Costs patient social care in state moderate dementia community setting | 0 | Idem | 653*12 | [table 2] |
| c.sev_sc | Costs patient social care in state severe dementia community setting | 0 | Idem | 1,095*12 | [table 2] |
| c.mci_i_sc | Costs patient social care in state MCI institution setting | 88,728 | [table E15] Costs per month multiplied by 12 | 8,762*12 | [table 2] |
| c.mil_i_sc | Costs patient social care in state mild dementia institution setting | 88,728 | Idem | 8,762*12 | [table 2] |
| c.mod_i_sc | Costs patient social care in state moderate dementia institution setting | 88,728 | Idem | 8,762*12 | [table 2] |
| c.sev_i_sc | Costs patient social care in state severe dementia institution setting | 88,728 | Idem | 8,762*12 | [table 2] |
| c.mci_ic | Cost informal care in state MCI community setting | 26,877 + 337 | Informal care costs (69 hours/month * 12 months * 32.46 hourly wage) [table E16; page E12] plus patient productivity cost (20.4% working * 4.9% work reduction * 20 hours/week * 52 weeks * \$32.46 hourly wage) [page E11]. Patient productivity loss is ad- hoc headed under informal care. | 754*12 + 988*12 | [table 2] |
| c.mil_ic | Cost informal care in state mild dementia community setting | 44,016 + 325 | Idem-MCI | 781*12 + 2,184*12 | [table 2] |
| c.mod_ic | Cost informal care in state moderate dementia community setting | 65,829 | Idem but with no patient productivity costs | 799*12 + 3,227*12 | [table 2] |
| c.sev_ic | Cost informal care in state severe dementia community setting | 116,077 | Idem-mod | 811*12 + 5,402*12 | [table 2] |
| c.mci_i_ic | Cost informal care in state MCI institution setting | 11,826 + 337 | [page E12] 44% of informal care in state community setting plus assumed same patient productivity loss as in state community setting | 754*12 + 435*12 | [table 2] |
| c.mil_i_ic | Cost informal care in state mild dementia institution setting | 19,367 + 325 | Idem | 781*12 + 961*12 | [table 2] |
| c.mod_i_ic | Cost informal care in state moderate dementia institution setting | 28,965 | Idem | 799*12 + 1,420*12 | [table 2] |
| c.sev_i_ic | Cost informal care in state severe dementia institution setting | 51,074 | Idem | 811*12 + 2,377*12 | [table 2] |
| c.Tx | Cost of treatment | 26,500 + 2,037 | Drug annual wholesale acquisition cost (\$26,500) [table E13] plus 2-weekly [table E12] treatment administration cost (\$78.35) [page E10] converted to year | 0 | Treatment costs were not part of research question |
| c.Tx_start | Additional costs in first cycle (not half-cycle corrected) | 1,044 + 168 | First-year monitoring MRI scan (\$261.10) [page E10] multiplied by 4 to reflect a scan every 3 months [page E10] plus 3 [page E10] adverse event ARIA MRI scans (\$261.10) [page E10] for 21.5% Probability of any ARIA [table E8] | $212.14 * 5 + 0.126 * 0.78 * 212.14 * 2 + 0.126 * 0.22 * 0.91 * 796.80 + 0.126 * 0.22 * (1-0.91) * 1098.27$ | [page 803; table 2] Monitoring (5x MRI in year 1) and ARIA-E (12.6%) being asymptomatic (78%) (2x MRI), symptomatic (22%) mild/moderate (91%) or symptomatic severe (9%) |
| discount_EFFECT | Discount rate for effects | 0.03 | [page 23] | 0.03 | [page 798] |
| discount_QALY | Discount rate for QALYs | 0.03 | [page 23] | 0.03 | [page 798] |
| discount_COST | Discount rate for costs | 0.03 | [page 23] | 0.03 | [page 798] |
| wtp | Willingness to pay threshold | 100,000 | Not used in base case results, set to ad-hoc value | 100,000 | Same as ICER |
| half_cycle_correction | Apply half-cycle correction | TRUE | We assumed this was applied but no description of half-cycle correction was found in the ICER report | TRUE | Assumption |

\* Table and page numbers refer to the original publications.

**Table S2:** Proportion in states over a 10-year time period and cumulative person-years in (aggregated care setting) states of ICER and AD-ACE cross-validation in IPECAD (undiscounted, not half-cycle corrected).

|  | ICER<br>IPECAD<br>cross-validation |  |  |  |  | AD-ADCE<br>IPECAD<br>cross-validation |  |  |  |  |
| --- | --- | --- | --- | --- | --- | --- | --- | --- | --- | --- |
| Cycle (year) | MCI | Mild | Moderate | Severe | Death | MCI | Mild | Moderate | Severe | Death |
| *Control*<br>*strategy* |  |  |  |  |  |  |  |  |  |  |
| 0 | 0.55 | 0.45 | 0 | 0 | 0 | 0.78 | 0.22 | 0 | 0 | 0 |
| 1 | 0.42 | 0.37 | 0.15 | 0.02 | 0.05 | 0.59 | 0.29 | 0.07 | 0.01 | 0.04 |
| 2 | 0.32 | 0.30 | 0.20 | 0.08 | 0.10 | 0.45 | 0.29 | 0.13 | 0.04 | 0.08 |
| 3 | 0.25 | 0.24 | 0.20 | 0.15 | 0.17 | 0.34 | 0.26 | 0.16 | 0.09 | 0.15 |
| 4 | 0.19 | 0.19 | 0.18 | 0.20 | 0.25 | 0.26 | 0.22 | 0.16 | 0.13 | 0.23 |
| 5 | 0.14 | 0.14 | 0.15 | 0.22 | 0.35 | 0.20 | 0.18 | 0.15 | 0.16 | 0.32 |
| 6 | 0.11 | 0.11 | 0.12 | 0.22 | 0.44 | 0.15 | 0.14 | 0.12 | 0.16 | 0.42 |
| 7 | 0.08 | 0.08 | 0.10 | 0.20 | 0.54 | 0.12 | 0.11 | 0.10 | 0.15 | 0.52 |
| 8 | 0.06 | 0.06 | 0.07 | 0.17 | 0.63 | 0.09 | 0.08 | 0.08 | 0.13 | 0.62 |
| 9 | 0.04 | 0.05 | 0.06 | 0.14 | 0.72 | 0.07 | 0.06 | 0.06 | 0.10 | 0.71 |
| 10 | 0.03 | 0.03 | 0.04 | 0.11 | 0.79 | 0.05 | 0.05 | 0.05 | 0.08 | 0.78 |
| Total | 2.19 | 2.02 | 1.27 | 1.51 | 4.04 | 3.10 | 1.90 | 1.08 | 1.05 | 3.87 |
| *Intervention*<br>*strategy* |  |  |  |  |  |  |  |  |  |  |
| 0 | 0.55 | 0.45 | 0 | 0 | 0 | 0.78 | 0.22 | 0 | 0 | 0 |
| 1 | 0.46 | 0.38 | 0.11 | 0.01 | 0.05 | 0.64 | 0.26 | 0.05 | 0.01 | 0.04 |
| 2 | 0.37 | 0.32 | 0.15 | 0.06 | 0.10 | 0.52 | 0.27 | 0.09 | 0.03 | 0.08 |
| 3 | 0.30 | 0.26 | 0.16 | 0.11 | 0.16 | 0.42 | 0.26 | 0.12 | 0.06 | 0.14 |
| 4 | 0.25 | 0.22 | 0.15 | 0.15 | 0.24 | 0.34 | 0.23 | 0.13 | 0.10 | 0.21 |
| 5 | 0.20 | 0.18 | 0.13 | 0.17 | 0.32 | 0.27 | 0.20 | 0.12 | 0.12 | 0.29 |
| 6 | 0.16 | 0.14 | 0.11 | 0.18 | 0.41 | 0.21 | 0.16 | 0.11 | 0.13 | 0.38 |
| 7 | 0.13 | 0.11 | 0.09 | 0.17 | 0.50 | 0.17 | 0.13 | 0.10 | 0.13 | 0.47 |
| 8 | 0.10 | 0.09 | 0.07 | 0.15 | 0.59 | 0.13 | 0.11 | 0.08 | 0.11 | 0.57 |
| 9 | 0.08 | 0.07 | 0.06 | 0.12 | 0.67 | 0.10 | 0.08 | 0.07 | 0.10 | 0.65 |
| 10 | 0.06 | 0.05 | 0.05 | 0.10 | 0.74 | 0.08 | 0.06 | 0.05 | 0.08 | 0.73 |
| Total | 2.66 | 2.27 | 1.08 | 1.22 | 3.78 | 3.66 | 1.98 | 0.92 | 0.87 | 3.56 |

**Table S3:**
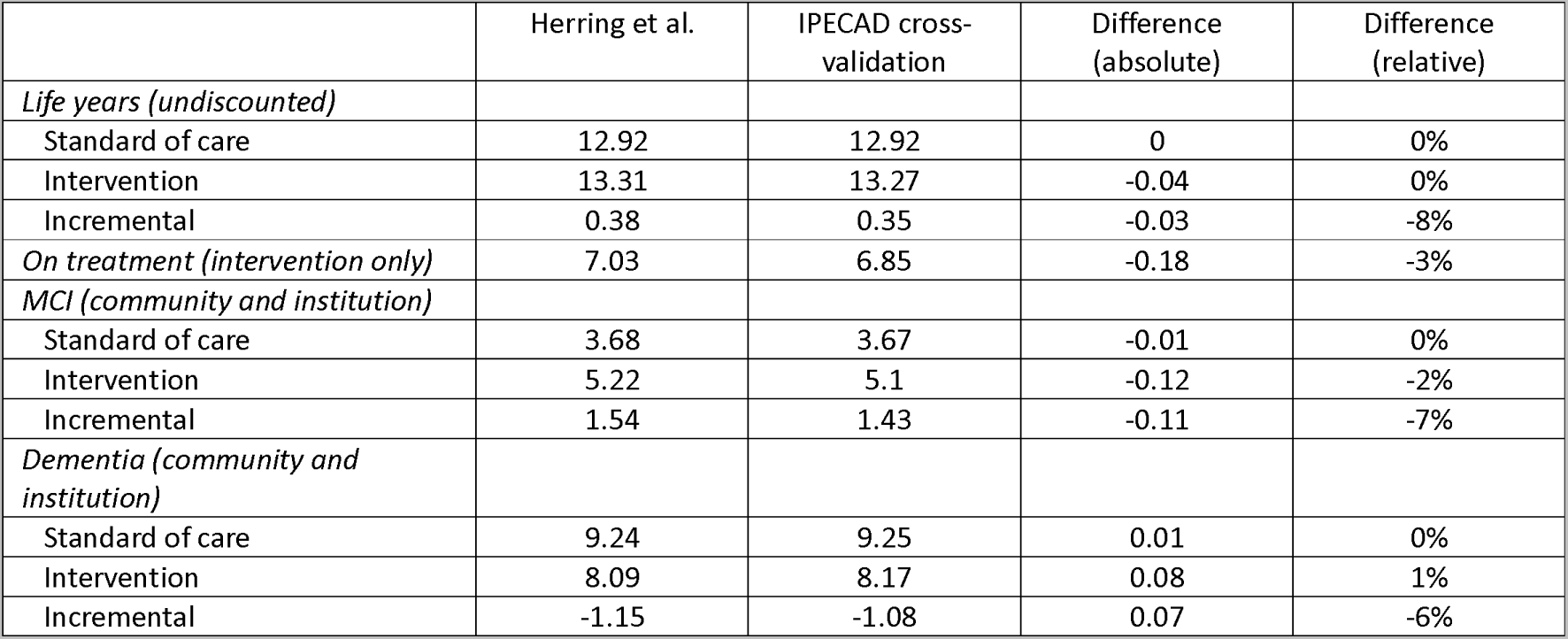
Cross-validation Herring et al. model; reported base case discounted model outcomes from Herring et al. and the IPECAD model outcomes from its cross-validation.

**Figure S1:**
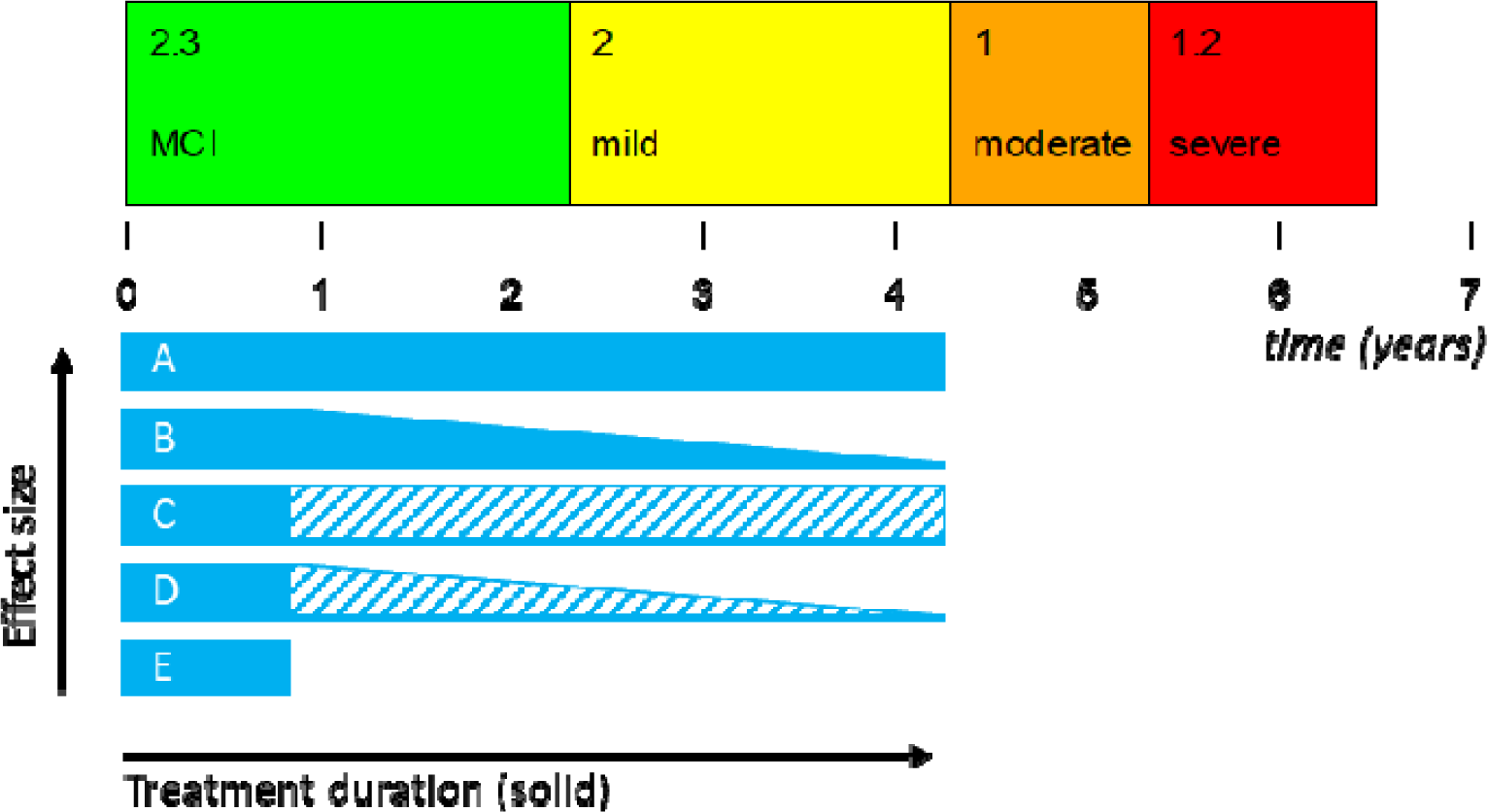
Graphical representation of assumptions on treatment discontinuation and effectiveness (waning) after the trial 18-month follow-up period (i.e., treatment extrapolation). A: Continue treatment & no treatment effect waning during treatment (base case) B: Continue treatment & treatment effect waning during treatment C: Stop treatment & treatment effect without effect waning after treatment stop D: Stop treatment & treatment effect with effect waning after treatment stop E: Stop treatment & no treatment effect after treatment stop

